# Service-user experiences of an integrated psychological intervention for depression or anxiety and tobacco smoking in IAPT: A qualitative investigation into mechanisms of change in quitting smoking

**DOI:** 10.1101/2022.03.23.22272703

**Authors:** Kim Fredman Stein, Katherine Sawyer, Shadi Daryan, Jennifer L Allen, Gemma Taylor

**Author notes:** **Declaration of Interest** Kim Fredman Stein, Jennifer Allen, Katherine Sawyer and Shadi Daryan have no conflicts of interest. Gemma Taylor has previously received funding from Pfizer, who manufacture smoking cessation products, for research unrelated to this study. **Funding source** Kim Fredman Stein was funded by Health Education England’s DClinPsy Training Programme during the conduct of this study. Gemma Taylor is funded by a Cancer Research UK Population Researcher Postdoctoral Fellowship award (reference: C56067/A21330) and Cancer Research UK project award (reference: PPRCPJT\100023). Katherine Sawyer is funded by Cancer Research UK Population Researcher Postdoctoral Fellowship award (reference: C56067/A21330). Shadi Daryan is funded by Cancer Research UK project award (reference: PPRCPJT\100023). Jennifer Allen does not have a funding source related to this study. **Ethical Approval** Ethics approval for this study was received from the NHS Research Ethics Committee on 12 July 2019 (Reference 19-245). Ethical approval for the IntEgrating Smoking Cessation treatment As part of routine Psychological care for dEpression and anxiety (ESCAPE) trial was obtained from a National Health Service Research Ethics Committee (19/03/2018, IRAS ID: 239339). **Author contribution** (using CREDIT guidance https://casrai.org/credit/) Kim Fredman Stein was involved in the following tasks: Conceptualization, Data curation, Formal Analysis, Investigation, Methodology, Project administration, Validation, Visualization, Writing – original draft, Writing – review & editing. Gemma Taylor was involved in the following tasks: Conceptualization, Data curation, Formal Analysis, Funding acquisition, Investigation, Methodology, Project administration, Resources, Supervision, Validation, Writing – review & editing. Jennifer Allen was involved in the following tasks: Supervision, Writing – review & editing. Katherine Sawyer was involved in the following tasks: Data curation, Formal Analysis, Investigation, Methodology, Project administration, Validation, Writing – review & editing. Shadi Daryan was involved in the following tasks: Project administration, Writing – review & editing. **Data Availability** Anonymised transcript data are available for free via application to the University of Bath’s Research Data Service https://www.bath.ac.uk/guides/research-data-service/.

## Abstract

**Background:** High smoking prevalence leads to increased morbidity and mortality in individuals with depression/anxiety. Integrated interventions targeting both smoking and mood have been found to be more effective than those targeting smoking alone, but the mechanisms of change of these integrated interventions have not been investigated.

**Aims:** This qualitative study aimed to investigate participants’ subjective experiences of the mechanisms underlying change in smoking behaviour following an integrated CBT-based intervention for smoking cessation and depression/anxiety.

**Methods:** This study was embedded within an ongoing randomised controlled acceptability and feasibility trial (http://www.isrctn.com/ISRCTN99531779). Semi-structured interviews were conducted with 15 IAPT service users and data were analysed using thematic analysis.

**Results:** Five themes were identified: (1) acquiring an increased awareness of smoking patterns, (2) developing individualised strategies, (3) practitioner style as “supportive but not lecture-y” (4) importance of regular sessions, and (5) having the opportunity to access intervention at “the right time”.

**Conclusions:** These findings further our understanding of the mechanisms of change towards smoking cessation in this integrated intervention and highlight the need to emphasise and embed these components in manualised interventions to optimise therapeutic benefits and reduce smoking prevalence in people with depression/anxiety.

**Relevance statement:** This study qualitatively investigated mechanisms underlying change in smoking behaviour following an integrated intervention for depression/anxiety and smoking cessation. Research shows interventions integrating smoking cessation and mood management are more effective than smoking cessation interventions alone; however further investigation is needed to understand mechanisms of change for integrated interventions. The current results identified key factors that were important for participants in the integrated intervention to reduce smoking. An improved understanding of mechanisms underlying change in smoking behaviour helps to identify therapist characteristics and treatment components that improve therapeutic outcomes which future research could investigate using a randomised controlled trial design.

## Introduction

Individuals with depression/anxiety are twice as likely to smoke than those without depression/anxiety (1), this disparity increases mortality in people with depression/anxiety compared to the general population [mortality rate ratio, 1.92 (95% CI: 1.91 to 1.94)](2). Despite being equally as motivated to quit, this population smoke more heavily, are more addicted, and are less likely to successfully quit than are the general population (3-5). There are many reasons for high smoking rates in this population, for example they are less likely to be prescribed smoking medicines (6), and it is widely believed that smoking cessation can worsen mental health (7, 8). However, a recent Cochrane review found evidence that smoking cessation can improve anxiety/depression compared to continuing smoking (standardised mean difference, -0.31 (95% CI: -0.40 to -0.22)) (9). Another Cochrane review found smoking cessation interventions offered alongside mood management support led to higher cessation rates compared to smoking cessation interventions alone for people with depression [risk ratio 1.47, 95% CI [1.13, 1.92]] (10); indicating the importance of integrated interventions to improve smoking and mood outcomes. However, this review did not shed light on the mechanisms that led to behavioural change.

Integrated interventions may be more effective for quitting smoking for various reasons. For example, cognitive behavioural techniques (CBT) could alter unhelpful beliefs about the relationship between smoking and depression/anxiety (e.g., “smoking helps my mood”) (11), which could promote cessation and prevent relapse. Such an intervention could also promote alternative strategies for managing depression/anxiety to smoking (12). For example, behavioural activation aims to increase pleasurable activities (13). It is also possible that the therapeutic alliance in psychological interventions could help facilitate behaviour change (14, 15).

There are some evidence-based models that we can use to investigate mechanisms of behavioural change. The Capability, Opportunity, Motivation-Behaviour (COM-B model) (16) suggests that a person’s capability (i.e., a person’s physical and psychological capacity), opportunity (i.e., external factors that facilitate behaviour) and motivation are involved in behavioural change. The Smoking, Not smoking, Attempting to quit, Planning to quit model (SNAP)(17) suggests that smoking cessation involves moving through the four stages of 1) smoking, 2) attempting to quit, 3) planning to quit and 4) not smoking. The misattribution hypothesis suggests that smokers misattribute nicotine withdrawal symptoms of stress, or anxiety/depression, and perceive smoking alleviates symptoms of mental illness (18, 19).

Understanding mechanisms of behavioural change and how they fit into evidence-based frameworks could improve our understanding of the active intervention components and help identify therapist characteristics that optimise therapeutic benefits, potentially leading to more effective and streamlined interventions.

Addiction research has been criticised for excluding the patient’s view and focusing on intervention techniques rather than intervention mechanisms (15). Therefore as part of a wider trial (20) the current study used qualitative interviews following an integrated intervention to investigate individuals’ subjective experiences of the mechanisms underlying change in their smoking behaviour.

In this qualitative investigation we aimed to use evidence-based models of behaviour change and behavioural intervention development (16-19) to explore participant’s subjective experience of mechanisms of change in smoking cessation.

## Methods

### Design

Our study was pre-registered (https://osf.io/nfgu4/) and was part of an on-going RCT (ESCAPE, http://www.isrctn.com/ISRCTN99531779). The anonymised data are available to researchers via application to the University of Bath (https://www.bath.ac.uk/guides/research-data-service/). We followed COREQ reporting guidelines in writing this manuscript (21). The authors assert that all procedures contributing to this work comply with the ethical standards of the relevant national and institutional committees on human experimentation and with the Helsinki Declaration. All procedures were approved by an NHS Research Ethics Committee (19/03/2018, IRAS ID: 239339). We used qualitative in-depth interviews to explore participants perceptions of change (22).

### Participants

We approached 19 patients and conducted interviews with 15 who took part in the ESCAPE trial intervention arm who had attended 3 or more intervention sessions. Participants in the intervention arm received a CBT-based smoking cessation intervention that was integrated into routine IAPT care for depression/anxiety (9, 20) (Supplement A). Improving Access to Psychological Therapies (IAPT) is a primary care service in the UK National Health Service providing evidence-based psychological therapies for depression/anxiety. Trial inclusion criteria were self-reported daily smokers of at least one year, aged ≥ 18 years; diagnosis of depression and/or anxiety (clinician administered PHQ-9 score ≥□10, and/or GAD-7 ≥□8 score) and were about to start IAPT treatment. Individuals were excluded if they did not have capacity to give informed consent, or if they were pregnant or breastfeeding at trial entry.

### Procedure & recruitment

Recruitment procedures for the ESCAPE trial can be found in the trial protocol(https://osf.io/nfgu4/). Purposive sampling was used to recruit participants for follow-up interviews about the intervention. During 3- and 6-month telephone ESCAPE trial follow-ups, participants were asked if they would like to be interviewed about their experiences in the study and attempting to quit. We recruited participants until information power was reached (23). Information power is more suitable for pragmatic applied health research than data saturation. ‘Data saturation’ was originally developed for grounded theory analysis (23, 24). Participants gave oral consent prior to the interview, and again as an audio recorded consent statement.

### Interviews

The interview schedule (Supplement B) aimed to explore participants’ experiences of mechanisms of change. Interviews lasted 30-45 minutes and were embedded within a longer interview schedule, lasting no more than 60 minutes, that also investigated the acceptability of the intervention and trial procedures (the additional findings will be presented elsewhere). Interviews were conducted by KM and KS.

### Service-User Consultation

Persons with lived experience of depression, anxiety and tobacco addiction contributed towards design of the interview schedule, participant information sheets, and the debriefing process.

### Analysis

Data were transcribed using a University approved service. Fifty percent of the audio data were checked against the transcripts to ensure fidelity. The data were analysed using reflexive thematic analysis following the steps outlined by Braun and Clarke (25, 26). Reflexive thematic analysis was used as it is not tied to theoretical or epistemological approaches and can be used both inductively and deductively. A critical realist approach was taken; meaning was viewed as both socially constructed and relating to individuals’ experiential reality (27). Braun and Clarke’s (25) guidance for the six phases of thematic analysis were followed: 1) familiarisation with the data, 2) coding, 3) generating themes, 4) reviewing themes, 5) defining and naming themes 6) writing up. A combined inductive and deductive approach to coding was taken, whereby codes and themes were developed both from existing theory and the data. The framework of deductive codes was constructed based on the COM-B model (Supplement C, Table S1) which has shown good reliability for categorising components of behaviour change interventions (28). Coding was conducted manually. Participant IDs were used throughout, and any potentially identifying information was removed.

Once the data were coded, all relevant coded data extracts were collated and organised into potential themes and subthemes. A series of tables were developed to explore possible relationships between codes, themes and subthemes. Potential themes and subthemes were then reviewed, refined, and assessed according to the criteria of internal homogeneity and external heterogeneity (29). Analysis was viewed as an iterative process; the researcher at times returned to previous stages rather than following a rigidly linear process. A self-reflexive stance was used throughout data collection and analysis to increase awareness of and limit the impact of the researcher’s potential biases and assumptions.

## Results

Fifteen participants were recruited, (Table 1; Supplement D, Table S2). We identified five themes, and 12 subthemes (Table 2), and present them with illustrative quotes (Table 3).

**Table 1.** Participant Characteristics Summary

| Participant Characteristics | N |
| --- | --- |
| Gender |  |
| Men | 6 |
| Women | 9 |
| Race/ethnicity |  |
| White British | 15 |
| Highest level of education |  |
| Higher degree | 4 |
| Degree | 6 |
| A-Level equivalent | 1 |
| GCSEO-grade/equivalent | 1 |
| Apprenticeship | 1 |
| Other vocational | 2 |
| Age (mean) | 42.6 |
| Age (range) | 26-67 |

*N* = 15 total

**Table 2.**
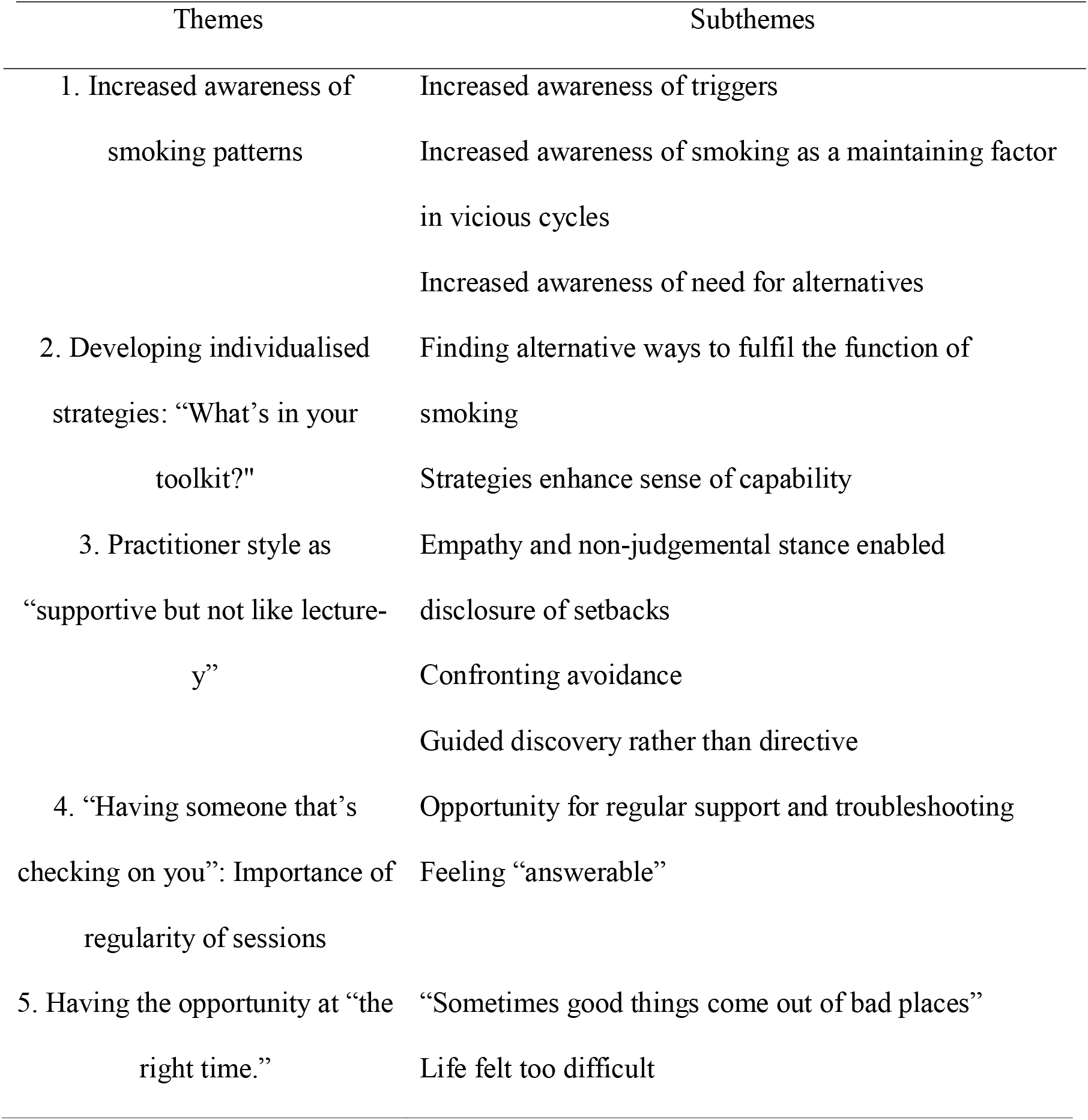
Themes and Subthemes

**Table 3.** Illustrative quotes from participants

| Theme | Subtheme | Quote(s) |
| --- | --- | --- |
| Theme 1:<br>Increased awareness of smoking patterns | Increased awareness of triggers | Quote 1: *“Once I started thinking a bit more about why I needed to do that, or why I was doing it, and if you're a bit more conscious of that element you can almost pre-empt it and divert it”* (Participant 4).<br>Quote 2: *“being aware of how you're spending your time and what is causing you to go, Oh, I need a cigarette (...). It was mapping out your day and working out when the trigger events were”* (Participant 14). |
|  | Increased awareness of need for alternatives |  |
|  | Increased awareness of smoking as a maintaining factor in vicious cycles | Quote 3: *“It was like a coping mechanism that, in the short term, seemed to help, but in the long- term then I'd worry about it (...), it was making me feel worse”* (Participant 3).<br>Quote 4: *“You have the cigarette, you feel a bit better but when the nicotine is less effective (...) you feel worse again and you have the cravings and you feel more stressed and more anxious again after it's worn off, which I never really thought about before”* (Participant 5).<br>Quote 5: *“I was conscious of my weight and thought, “If I smoke, I'm going to have less of an appetite” (...) I'm a healthy weight and the way I was thinking was, and acting when I was younger was making me underweight and unhealthy, and I think it was just like having a realisation of that”* (Participant 13). |
| Theme 2:<br>Developing individualised strategies: “What's in your toolkit?” | Finding alternative ways to fulfil the function of smoking | Quote 6: *“I'd be anxious when I came home from school and I still had a lot of work to do, and I'd have a wine and sit in the courtyard and smoke because that helped, but in the short term. So, we looked at what could you do differently, so could you go for a run, could you go for a swim, could you play the piano, and things like that. So, then it gave me a bit more, changed the behaviour, rather than just stopped the smoking”* (Participant 3). |
|  | Strategies enhance sense of capability | Quote 7: *“the strategies are really helpful- rather than just going, ‘OK, I just will stop this habit I've had since I've been, 21 years’, that's a bit impossible, well, I found it a bit impossible before”*. (Participant 3).<br>Quote 8: *“you could see that you were doing better, when they did the carbon monoxide test”* (Participant 1).<br>Quote 9: *“it was like, ‘wow, look how much money I've saved but also how much time I've saved’”* (Participant 3).<br>Quote 10: *“when I had the Champix, because it made the actual smoking make me feel sick, you couldn't smoke, it completely put you off it”* (Participant 2).<br>Quote 11: *“Because I wasn't really craving it because I sort of had the nicotine, but yeah, it was a lot easier and easier to manage at work. I was calmer, not trying to give up and work and get stressed, so that was really helpful”* (Participant 1).<br>Quote 12: *“I would not smoke for a couple of days but then I'd just find* |
|  |  | *myself getting drawn back into doing it, through having colleagues that smoked or friends that smoked when we were out” (Participant 13).*<br>Quote 13: *“then they’re going to be aware of me doing it, and not be likely to offer me a cigarette (...) it did work and I think it did help having that” (Participant 13).* |
| Theme 3:<br>Practitioner style as “supportive but not like lecture-y” | Empathy and non-judgemental stance enabled disclosure of setbacks | Quote 14: *“If you’ve got somebody that you’re not connecting with (...) And that doesn’t seem to be understanding of you, you’re not going to take their advice because it’s going to be: you don’t understand where I’m coming from, how can you expect me to do something when you don’t understand? Showing no empathy for what’s going on. It makes you put a wall up, I suppose, and you get quite defensive. Whereas if you’ve got somebody that you get on with, you can open up to, that seems to understand what you’re going through, the wall comes down so you’re more likely to take on-board what’s said” (Participant 2).*<br>Quote 15: *“she was very understanding of what I was going through as a whole anyway but it just meant that when it came to the smoking bit, it helped make that a more comfortable environment to discuss it” (Participant 13).*<br>Quote 16: *“The temptation is to be very self-critical when something you try doesn’t work, and actually having somebody say, “It doesn’t matter, these things happen, it’s absolutely fine for it not to have worked this time, let’s just have a reboot”, something about what it was that caused the hiccup last time, “See if we can try and avoid that next time” (...) There was no judgement or condemnation” (Participant 4).*<br>Quote 17: *“as soon as I turned around and said, “Yeah, I haven’t had one” she said, “Oh that’s fantastic!” Even just that little bit of encouragement was really good” (Participant 5).* |
|  | Confronting avoidance | Quote 18: *“She wasn’t avoiding talking to me about, which was brilliant, because, “Let’s talk about the smoking”, so I knew I had to confront it, it wasn’t, “You said you’d stop smoking last week and you didn’t so we’re just going to ignore it”, it was, “What happened this time that made you feel...?”, so it was examining it, which is great” (Participant 6).* |
|  | Guided discovery rather than directive | Quote 19: *“I feel it was subtly getting me to make the decisions and getting me to make the choices (...) it was very much guiding rather than leading” (Participant 4).*<br>Quote 20: *“I guess the only thing I would suggest is if they had a bit more time for calls so they didn’t feel ... because you know that they’re getting stressed too. The person who’s supposed to be counselling you is getting stressed because they’ve got to get onto the next thing, and you can sense that they’re trying to interpret what you’re saying so they can get onto the next because they have six more questions and there’s only four minutes left” (Participant 10).* |
| Theme 4:<br>“Having someone that’s checking on you”: Importance of regularity of sessions | Opportunity for regular and frequent support and troubleshooting | Quote 21: *“if there was a blip, which there were, they didn’t fester for too long before I was able to talk about it or get some more advice” (Participant 4).*<br>Quote 22: *“knowing you’re going to phone and speak to someone every week about it made it that bit easier for me to carry on and do” (Participant 5)*<br>Quote 23: *“It was the fact that I had to report into someone and look into why, that was what helped me rather than, so I think I didn’t do it before* |
|  |  | *because I just felt like it was too big a job to do on my own*” (Participant 3). |
|  | Feeling “answerable” | Quote 24: *“having somebody checking on you, you’ve got to tell them that you’ve smoked, and you feel like you’ve let somebody else down rather than just yourself”* (Participant 7). |
| Theme 5: Having the opportunity at “the right time.” | “Sometimes good things come out of bad places” | Quote 25: *“if I’m going to change to my whole mental state, I might as well try and change everything at the same time, and it just seemed a perfect opportunity to give it a go”* (Participant 8). |
|  | Life felt too difficult | Quote 26: *“It just seemed to be going from one trauma to the next, and I was just running out of energy to deal with everything, and quitting smoking just became very low on my list of priorities (...) I don’t think anything would have helped, because I had to deal with the issues I was dealing with, and till I’d dealt with those I couldn’t contemplate anything else”* (Participant 2).<br>Quote 27: *“I’ve still got the patches because I didn’t use them all, so I have got some backup almost if say tomorrow I go, “That’s it”, at least I’ve got that (...), it’s almost like I’m in constant preparation because they’re there, so that’s good”* (Participant 6). |

### 1. Increased Awareness of Smoking Patterns

Participants described an increased awareness of smoking patterns as a key step in facilitating quitting smoking. They described a shift in their awareness, from smoking being automatic where they would “smoke and not necessarily know I was smoking” (Participant 2), to being more “analytical” about their smoking patterns (Participant 4). This “understanding why you smoke” (Participant 3) seemed to facilitate participants’ sense of psychological capability to not smoke (Table 3, Quote 1).

One aspect of increased awareness was an improved awareness of smoking triggers (Table 3, Quote 2). One of the most common triggers for smoking was strong negative emotions. Participants expressed through the intervention, they gained increased awareness of the link between strong negative emotions and smoking: “when I talk about it, I do smoke because I’m unhappy and because it’s a distraction” (Participant 11). Some expressed the realisation that smoking was often an attempt to cope with negative emotions: “before, I felt that if I was stressed, I could step away and have a fag and that would help” (Participant 5). This response reflects the misattribution hypothesis (18). The increased awareness of this misattribution seemed to result in a change in thinking about the relationship between smoking and stress, as well as providing alternative options for managing stress: “I recognised that and that’s more just stepping out of the situation and just chilling for a bit” (Participant 5). Therefore, increased awareness of the need for alternatives to smoking for managing emotions appears to be a mechanism of change in reducing smoking.

Participants also described greater awareness of how smoking contributed to the maintenance of a “vicious cycle” (Participant 13) of smoking and stress (Table 3, Quote 3). Several participants identified an increased awareness of the specific role of nicotine withdrawal in maintaining unpleasant feelings (Table 3, Quote 4).

This increased awareness of how smoking was contributing to difficulties in their mental health appeared to increase motivation to change. One participant described smoking to control her appetite; increased awareness of the unhelpful nature of her beliefs about her weight helped to facilitate stopping smoking (Table 3, Quote 5). This suggests that there are a range of beliefs which drive smoking, and an increased awareness of these allows them to be challenged. Moreover, increased awareness that smoking appears to help with strong negative emotions in the short-term, but maintains them in the long-term, seemed to increase motivation to quit smoking, and understanding of the need to develop other ways to cope with emotions.

### 2. Developing Individualised Strategies: “What’s in Your Toolkit?”

All participants described developing a personalised set of strategies that they found helpful in reducing smoking, described as acquiring “a couple of tricks up your sleeve” (Participant 10). These strategies included goal setting, setting a quit date, problem-solving, removing access to cigarettes, communicating with others about quitting, positive self-talk, distraction, breathing and mindfulness techniques, and using smoking cessation resources such as apps. It seemed particularly important to develop “substitutes” (Participant 12) which fulfilled the function of smoking (Table 3, Quote 6).

Practical strategies provided by the intervention appeared to make the quit attempt feel more manageable and enhance participants’ sense of psychological capability (Table 3, Quote 7). Similarly, Participant 13 specifically referred to “exercises for breathing and mindfulness and distraction techniques” that helped her to feel capable: “No, I don’t have to have a cigarette (…) I’m stronger, I can deal with that”. Several strategies also enhanced motivation; namely, goal setting, setting a quit date, and using a pros and cons list which helped to “cement” motivation for change (Participant 4). Once participants noticed concrete signs of improvement resulting from these strategies, this reinforced their motivation to continue to implement them, as well as enhancing their sense of psychological capability (Table 3, Quote 8 & 9).

Although smoking cessation medication was not viewed as helpful by everyone, some participants found it to be an important tool (Table 3, Quote 10). It seemed particularly helpful for managing the physiological nicotine cravings, which enhanced some participants’ sense of their physical capability to quit (Table 3, Quote 11).

The strategy of communicating with others about quitting was viewed as important by several participants. They described being with others who were smoking as a trigger (Table 3, Quote 12). However, telling others about the quit attempt also helped (Table 3, Quote 13). This suggests that changing the social environment can facilitate behaviour change in smoking, consistent with social learning theory principles (30).

### 3. Practitioner Style as “Supportive but not Like Lecture-y”

Participants expressed how important the therapeutic alliance was in making changes to their smoking behaviour, particularly their sense of having the space to talk through their feelings and difficulties, with these being heard and not judged but met with compassion, curiosity, and support. This contrasted with descriptions of other people in their lives who tended to take a more judgemental and directive approach towards their smoking and was often experienced by participants as unhelpful: “lecture doesn’t have any impact” (Participant 6). The IAPT practitioners’ understanding, and empathy seemed to enable participants’ disclosure of when they had struggled with their quit attempt one week, as well as participants’ openness and receptiveness to suggestions (Table 3, Quote 14).

Additionally, participants expressed a sense that IAPT practitioners’ understanding of their mental health struggles helped them to feel more able to talk about smoking (Table 3, Quote 15). One participant expressed that the practitioner’s non-judgemental approach to setbacks was particularly important given the participants’ tendency to be self-critical (Table 3, Quote 16).

This non-judgemental IAPT practitioner stance enabled participants to learn from setbacks that may otherwise have served as a trigger to give up on smoking cessation. Given that levels of self-criticism are often high in depression (31), and that setbacks could be experienced as “an added failure” (Participant 15), this therapeutic stance is likely to be particularly important in this population. Alongside viewing setbacks as an opportunity for learning, IAPT practitioners’ encouragement enhanced participants’ motivation to continue with their attempt to quit (Table 3, Quote 17).

Several participants found it helpful that the IAPT practitioner helped them to confront their avoidance of talking about smoking or attempting to quit (Table 3, Quote 18).

Participants also noted the IAPT practitioner use of guided discovery to empower them to think through decisions themselves (Table 3, Quote 19). Sometimes practitioner empowerment of participant decision-making took more explicit forms, such as giving options around the use of smoking cessation medication.

One participant expressed that an unhelpful experience was that he felt rushed at times, rather than the IAPT practitioner responding to him in a flexible and person-centred way (Table 3, Quote 20).

### 4. “Having Someone that’s Checking on You”: Regularity of Sessions

Most participants expressed the importance of the regularity of sessions, which supported the development and maintenance of the therapeutic alliance. One helpful aspect of regular sessions was having the space to solve problems as they arose (Table 3, Quote 21). Regular check-ins seemed to enhance participants’ sense of psychological capability compared to when they had tried to quit on their own, due to the extra support and sense of accountability (Table 3, Quote 22 & 23).

Feeling “answerable” (Participant 4), or “responsible” (Participant 11) to the IAPT practitioner each week appeared to enhance participants’ motivation or even made them feel “obliged” (Participant 13) to quit (Table 3, Quote 24). This sense of anticipatory guilt seemed to drive reduced smoking for some participants. This sense of accountability was not limited to the IAPT practitioner; one participant recruited friends and family to have “more people to check in on how I was doing” (Participant 13).

### 5. Having the Opportunity at the “Right Time”

Several participants talked about the combination of feeling that it was “the right time” to change, and the integrated mental health intervention providing an opportunity, or the “push” (Participant 5) they “needed” (Participant 4). For these participants, mental health problems did not prevent it from being the right time: “although I was in a very bad place at that time, it was the right time” (Participant 4). In fact, beginning therapy for their mental health in some cases appeared to signal their readiness to change (Table 3, Quote 25). However, a couple of participants expressed that although they valued the opportunity, stressful life events made quitting seem too much (Table 3, Quote 26). For another participant, it appeared that the intervention helped her to approach “the right time”, even though she was not there yet. This suggests that the intervention helped her shift from attempting to quit to planning to quit, a shift consistent with the SNAP model of smoking cessation (17) (Table 3, Quote 27). Thus, having the opportunity for support with smoking cessation enhanced her motivation “because it was offered (…) I probably was more motivated to do it, have a go” (Participant 6).

## Discussion

### Summary of findings

In this qualitative study, we investigated participants’ subjective experiences of mechanisms underlying change in smoking behaviour. Participants reported that the integrated smoking cessation and mood intervention helped facilitate reductions in smoking through increased self-awareness of smoking patterns, and supported them to develop individualised behavioural and cognitive strategies to aid cessation. Participants viewed the regularity of support, the supportive, “non-lecture-y” therapeutic style of IAPT practitioners, and being offered the smoking intervention at the “right time” (i.e. integrated with mental health support) all contributed to participants’ sense of being able to make changes to the smoking. These findings further our understanding of the active ingredients and processes for behaviour change in this integrated intervention.

### Study Strengths and Limitations

Participants were all white British; this limits the study’s transferability to people of different backgrounds. Furthermore, lack of a longer-term follow-up means that we could not show whether intervention mechanisms were maintained long-term. When interpreting qualitative results, it is important to acknowledge the researcher’s background (32); KFS is a non-smoker, whose primary orientation clinically is CBT. These factors are likely to have influenced the interviews and analysis, although research supervision was used reflexively throughout the process to consider the researcher’s position and reflect on alternative perspectives.

This study possesses considerable strengths. According to Malterud’s et al. (23) criteria for information power, the study had strong information power as the sample was very specific to the research question and the study had clear theoretical underpinnings. Moreover, including people with lived experience in research design, including the interview schedule, to help ensure that questions were acceptable, relevant and non-judgemental for those who did not manage to quit smoking. Past studies have shown that integrated interventions can be effective but have not explored how and why they work (10). Our qualitative approach shed light on how participants made sense of what helped them reduce their smoking, situating these perceived mechanisms within the context of the participants’ lives.

### Comparison with existing literature

#### Increased Awareness of Smoking Patterns

This theme reflects one mechanism underlying several behaviour change techniques identified in Michie et al.’s (33) taxonomy of behaviour change techniques for smoking cessation, such as prompting self-recording, facilitating an understanding of how lapses occur, and identifying barriers to change. The current study improves our understanding of why these techniques can facilitate reductions in smoking. First, increasing awareness of the function of smoking for an individual can facilitate awareness of alternative ways of fulfilling this function. Second, increasing awareness of how smoking can contribute to a vicious cycle which worsens mental health by maintaining difficulties such as stress (through nicotine withdrawal), or worries about health in the long term, appeared to increase motivation to change. Participants expressed how the experience of smoking relieving the unpleasant symptoms of nicotine withdrawal can become overgeneralised, leading to the misattribution that smoking can relieve stress, consistent with the misattribution hypothesis (9, 18, 19). Increased awareness of this resulted in a shift towards viewing smoking as exacerbating stress rather than relieving it, leading to an awareness of the need for alternative ways to cope with unpleasant emotions. Third, there were a range of beliefs that may drive smoking (including weight concerns), and that increasing awareness of these beliefs is an important first step towards addressing them. This theme is consistent with traditional CBT theory, which emphasises the role of increasing awareness of triggers and maintenance cycles (34), and third-wave CBT approaches such as Mindfulness-based Cognitive Therapy which highlight the centrality of awareness in behaviour change (35).

#### Developing individualised strategies: “What’s in your toolkit?”

Most strategies mentioned by participants are included in Michie et al.’s (33) taxonomy of behaviour change techniques in smoking cessation interventions. However, some participants also found additional techniques, such as breathing and mindfulness helpful. This suggests that incorporating some “third wave techniques” into standard CBT for smoking cessation may also be helpful. This theme highlights why the inclusion of practical, individualised strategies is important. First, they help participants to find alternative ways to fulfil the function of smoking (e.g., relaxation), consistent with behavioural theory (36). Second, strategies can enhance participants’ sense of psychological capability to achieve the desired change in behaviour. Third, once participants found individualised strategies that were effective, this provided an experience of success. This reinforced their use of the technique and increased their motivation to continue with the quit attempt.

#### Practitioner style as “Supportive but Not Like Lecture-y”

Therapeutic style was an important factor in facilitating perceived smoking behaviour change. Rapport may have been enhanced as practitioners delivering the integrated intervention were experienced in promoting a therapeutic environment. Building “general rapport” is included as an intervention component in Michie et al.’s (33) taxonomy of behaviour change techniques. Results highlight how therapeutic style influences perceived treatment outcomes by enhancing or diminishing participants’ motivation and sense of capability to change. Consistent with a motivational interviewing stance, participant descriptions of helpful aspects of IAPT practitioner style included eliciting thoughts from the client rather than imposing their own views and empathic listening (37). Whereas one unhelpful aspect of practitioner style was the perception of being rushed, and of the practitioner paying more attention to manual pro-forma than to the participant. This indicates the importance of allowing sufficient time and flexibility in the implementation of the intervention and maintaining focus on the client’s needs during sessions.

The importance of the therapeutic alliance to promote behaviour change is consistent with past research showing that the strength of the therapeutic alliance predicted reduced cannabis use in young people at three and six-month follow-up assessments (14). Furthermore, in the UK Alcohol Treatment Trial, therapist characteristics and therapeutic alliance were the most common reasons given by participants for their successful reduction of alcohol use (15). This study therefore extends this finding to smoking cessation and is relevant given Orford’s criticism of addiction research as “neglecting relationships in favour of techniques” (38).

#### “Having Someone that’s Checking on You”: Importance of Regularity of Sessions

This theme highlights the importance of providing the opportunity for regular IAPT practitioner check-ins and encouraging individuals to ask others in their network to check-in on them, to enhance the sense of feeling “answerable”. This suggests that interventions should aim to keep sessions regular and could explore recruiting social networks for check-ins with clients. The importance of feeling “answerable” is consistent with the literature on the role of social processes in addiction mutual help organisations such as Alcoholics Anonymous (39).

#### Having the Opportunity at “the Right Time”

Participant ability to quit smoking coinciding with the opportunity to access support for smoking cessation whilst receiving mental health therapy is consistent with the COM-B model (16). The presence of mental health problems did not prevent participants from feeling ready to make changes to their smoking. This is important because individuals with mental health problems are not given the same opportunities for smoking cessation support as the general population (40), and there is a widely held belief among healthcare professionals that smoking cessation should only be attempted after mental health has improved (7, 8). These findings reinforce previous research suggesting that individuals with depression or anxiety can be motivated and supported to successfully reduce or quit smoking when given the opportunity (10, 41). However, this did not apply to all participants, with some expressing that life felt too difficult to quit. Nevertheless, the intervention appeared to help some participants move closer to “the right time”, into the “preparing to quit” stage of smoking cessation (17), suggesting the intervention may assist individuals to progress in their change process.

### Research and practice implications

These findings further our understanding of the active ingredients and processes for behaviour change in integrated smoking and mental health interventions. In terms of clinical practice, we have outlined the strategies and the therapeutic stance that could be embedded and emphasised in manualised interventions to optimise therapeutic benefits. Findings indicate that practitioners should not assume that having anxiety or depression means that individuals are not ready or motivated to quit smoking. These processes are currently being implemented in the ESCAPE trial, however, a full-scale trial is required to investigate the effectiveness of these components. Future research in this area should strive to recruit people from diverse cultural backgrounds and include a longer-term follow-up.

## Conclusions

Several key factors were identified by participants to be important in quitting smoking: gaining an increased awareness of smoking patterns; developing individualised strategies; having a practitioner with an empathic, non-judgemental and motivational stance; regular sessions to promote a sense of accountability and solve problems as they arise; and being given the opportunity at a time when individuals feel ready to change. If similar results are found in more diverse samples, these aspects should be embedded and emphasised within practitioner training and integrated interventions for smoking cessation and depression/anxiety. Future research could use the current findings to inform the constructs that should be targeted for study in research exploring mechanisms of change within a randomised controlled trial design.

## Data Availability

Anonymised transcript data are available for free via application to the University of Baths Research Data Service

https://www.bath.ac.uk/guides/research-data-service/.

## Acknowledgements

We would like to thank the participants in this research and gratefully acknowledge CRUK for funding this research.

