## Supplement for "Service-user experiences of an integrated psychological intervention for depression or anxiety and tobacco smoking in IAPT: A qualitative investigation into mechanisms of change in quitting smoking"

**Online supplement**

### Supplement A: Overview of Intervention Components

| **IAPT treatment appointment** | **1** | **2** | **3-5** | **6** |
| --- | --- | --- | --- | --- |
| **Smoking cessation session** | **Pre-quit** | **Quit day** | **Follow-up** | **Final** |
| Address beliefs about smoking and mental health | ✔ | ✔ | ✔ | ✔ |
| Inform the client about the treatment programme | ✔ |  |  |  |
| Assess current smoking | ✔ |  |  |  |
| Assess past quit attempts | ✔ |  |  |  |
| Explain how tobacco dependence develops and assess nicotine dependence | ✔ |  |  |  |
| Explain the importance of abrupt cessation and the ‘not a puff’ rule | ✔ | ✔ | ✔ | ✔ |
| Inform the client about withdrawal symptoms | ✔ |  |  |  |
| Discuss stop smoking medications | ✔ |  |  |  |
| Set the Quit Date | ✔ |  |  |  |
| Prompt a commitment from the client | ✔ | ✔ |  |  |
| Check on client’s progress |  |  | ✔ | ✔ |
| Confirm readiness and ability to quit |  | ✔ |  |  |
| Confirm that the client has a sufficient supply of medication |  | ✔ | ✔ | ✔ |
| Enquire about medication use |  |  | ✔ | ✔ |
| Discuss withdrawal symptoms and cravings, and how to cope |  | ✔ | ✔ |  |
| Advise on changing routine |  | ✔ |  |  |
| Discuss how to address the issue of the client’s smoking contacts and how the client can get support during their quit attempt |  | ✔ |  |  |
| Discuss any difficult situations experienced and methods of coping |  |  | ✔ | ✔ |
| Address any potential high-risk situations in the coming week |  | ✔ | ✔ |  |
| Discuss plans and provide a summary | ✔ | ✔ | ✔ | ✔ |
| Medication (NRT or Varenicline) | ✔ | ✔ | ✔ | ✔ |

### Supplement B: Interview Schedule

**Questions about recruitment/sign-up**

Can you tell me a little about how you found out about the study?

Can you tell me a little about why you decided to sign up?

Can you tell me a little about the treatment you received?

**Questions about quitting**

How was your experience of attempting to quit?

What aspects of the quit attempt did you find difficult?

Were there any aspects that you found easy?

What helped or got in the way of you quitting smoking?

What did you like/not like about the smoking cessation support you received?

Was there anything you would change about the smoking cessation support?

How did you feel when you were trying to quit/quitting/relapsing?

If relapsed…

What were the triggers?

Would there have been anything that could have been done differently in the intervention that you feel would have helped you to avoid relapsing?

Did you feel that your quit attempt impacted on your mental health?

Prompts: In the first few weeks compared to later on?

How did you find having smoking cessation support alongside therapy for your mental health?

How did you feel the smoking cessation support fitted alongside your psychological therapy?

Was there anything your therapist did particularly well?

Was there anything your therapist could have done better?

Would you recommend the intervention to a friend, if so, why?

**Questions about Mechanisms**

What did you think about the impact of smoking on your mental health before the intervention?

Do you feel that the intervention changed how you think about smoking and mental health?

Do you feel that the intervention changed what you do in relation to smoking?

Prompt: for example, the way you plan for situations in which are likely to trigger desire to smoke

Did you feel that your therapist was supportive in helping you to quit?

What did you like/not like about the smoking cessation support from your therapist?

How important was your relationship with your therapist in helping you to make changes to your smoking?

Have you had previous opportunities to access smoking cessation support?

(If yes) what was different about previous support? What was similar?

How motivated were you to quit before the intervention? (out of 10?)

How motivated were you to quit after the intervention? (out of 10?)

What were your motivations to quit? Did they change during the intervention?

What were your motivations to continue smoking? Did these change during the intervention?

Before beginning the intervention, how capable did you feel of quitting? What were the obstacles?

Did this change during the intervention?

(If there was a change) – What was it about the intervention that helped you to feel more capable of quitting?

Was there anything else that we haven’t already talked about that would be helpful for us to know about your experience of the therapy?

### Supplement C: Table S1 Deductive codes

| **Capability** | **Opportunity** | **Motivation** |
| --- | --- | --- |
| Physical | Social | Automatic (processes involving impulses and emotions) |
| Psychological | Physical | Reflective (i.e., plans and evaluations) |

### Supplement D: Table S2 Individual participant characteristics

| **ID** | **Age group** | **Sex** | **Highest level of education** |
| --- | --- | --- | --- |
| 1 | 31-40 | Female | Degree |
| 2 | 41-50 | Female | Apprenticeship |
| 3 | 31-40 | Female | Other vocational |
| 4 | 51-60 | Female | Degree |
| 5 | 21-30 | Male | Degree |
| 6 | 51-60 | Female | Degree |
| 7 | 31-40 | Female | Higher degree |
| 8 | 51-60 | Male | Higher degree |
| 9 | 31-40 | Male | GCSE/O-grade/equivalent |
| 10 | 61-70 | Male | Degree |
| 11 | 41-50 | Female | Other vocational |
| 12 | 41-50 | Male | Higher degree |
| 13 | 21-30 | Female | Degree |
| 14 | 41-50 | Male | Higher degree |
| 15 | 61-70 | Female | A-level equivalent |
